# A Scoping Review: How should fractures of the Anterior Process of the Calcaneus be managed in athletes?

**DOI:** 10.1101/2024.10.18.24315738

**Authors:** Joseph Franklin, Charlie Offer, Lucy Hammond, Hollie White, Samantha Wilson-Thain

## Abstract

The Anterior Process of the Calcaneus is part of the “heel bone” (calcaneus), located near to where this bone meets the Navicular bone. Both bones are in the foot. The main mechanism that causes this injury, in the sporting context, is an avulsion injury. Essentially, a ligament, that attaches the calcaneus to the navicular, is pulled with such force that it pulls off some of the attached calcaneus bone. Commonly this occurs when athletes “roll” their ankles.

Historically, these fractures have often been misdiagnosed, with fractures being mistaken for bad “sprains” and soft tissue damage. This misdiagnosis has led to the mismanagement of these injuries with poor patient outcomes and recovery. Despite growing amounts of guidance available, there is still a lack of clarity in the literature. This is particularly the case in how this injury should be managed in the athlete looking to return to sport.

This review seeks to analyse the preexisting guidance in the management of these injuries, pulling resources together, to better direct the clinician and improve patient/athlete outcomes. This review will also highlight areas where more research is needed.

## Background

The Anterior Process of the Calcaneus is part of the “heel bone” (calcaneus), near to where this bone meets the Navicular bone. A fracture of the anterior process (AP) has two main mechanisms. The first and the most popular is an avulsion fracture. It occurs commonly in a sporting context when an athlete ‘rolls’ their ankle. It is caused by an inversion motion of the plantar-flexed ankle with subsequent avulsion of a fragment of bone from the anterior process of the calcaneus. The second mechanism is a dorsiflexion fracture. It is caused by a forced dorsiflexion motion with the foot in eversion, with subsequent compression of the anterior process of the calcaneus between the cuboid and the talus. It is thought to occur predominantly in females due to the use of high-heeled shoes. AP fractures are commonly classified using the Degan system which is divided into three categories: Type 1, 2 and 3.

Historically, there have been high rates of misdiagnosis or delayed diagnosis of AP calcaneus fractures. The injury is rare (2–5% of all ankle sprains are associated with a fracture to the AP) and has relatively little medical research on the subject (Baljinder, 2019). AP fractures can be confused with injuries to the anterior talofibular ligament (AFTL) due to the close anatomical relationship between the pair. It is thought that up to 88% of AP fractures are missed on plane radiographs (Schepers, 2011).

Currently there is little consensus within the medical community on the best way to treat and manage these injuries. Current treatment approaches are split between non-operative treatment and surgical treatment. The former ranges from full weight-bearing to immobilization and non-weight bearing for a period of up to 10 weeks. Surgical treatment includes open reduction and internal fixation or open arthroscopic excision. Baljinder (2019) suggested that minimally displaced fractures should be managed initially non-operatively and that acute surgical intervention could be indicated for large type III fractures. Trnka (1998) stated that Type III fractures can cause cartilage lesions and arthrosis. Consequently, quicker treatment is needed, generally via excision or refixation of the fragment. Massen (2019) suggests that functional treatment of fractures through full weight-bearing produced positive results and a fast return to work for most patients. However, a prolonged period for the return to sporting activity was noted.

This review seeks to analyse and summarise the preexisting literature in the management of these injuries, to make recommendations as to how full function can be regained after injury, thus allowing athletes to return to sport. Due to the relative lack of literature on the subject we will also suggest avenues for potential future studies to address the gaps in knowledge and ultimately improve the outcome for patients.

### Aims & Objectives

The aim of this Scoping Review is to identify what the current literature tells clinicians on the management of fractures of the Anterior Process of the Calcaneus. The focus will be identifying management options that allow the patient to return to full function, as this is the outcome that will be needed by athletes seeking to return to sport, with minimal loss to performance. It is also important briefly evaluate the mechanism of injury, presenting symptoms, and investigation findings, to explore why these injuries are so often misdiagnosed, as this plays a large role in the management. The options available in managing these fractures will then be analysed, with the aim of providing clear guidance on how these injuries can be best treated and rehabilitated to allow patients to return to sport.

To meet these aims, a thorough understanding and evaluation of the current literature must be collated and reviewed. It is predicted that guidance on the management will not be unanimous or underpinned by sufficient research. Therefore, this Scoping Review will also aim to direct how future studies could be best directed to help provide further clarity for clinicians to rule out ambiguity.

### Project approach, methods & analysis

A Scoping review will be used to provide clearer guidance for clinicians to manage Anterior Process of the Calcaneus fractures. At present, there are not enough Randomised Control Studies to enable a Systematic Review in the management of these injuries. As such, a Scoping Review is well placed to amalgamate the available literature and direct research within this area.

There are numerous guides available as to how researchers should conduct a Scoping Review, which is still a relatively new tool. For this project we have decided to use the framework proposed by Mak and Thomas (2022). Located on the National Library of Medicine, it is a well cited and current paper that builds on the foundations of previous works within this area. The steps that Mak and Thomas outline for conducting Scoping Reviews are as follows:

1. Identify a Team
2. Identify the Research Question
3. Identify Relevant Studies
4. Select Studies to be included in the review
5. Chart the data
6. Collate, summarise and report the results
7. Consult the stakeholders (optional)

Using and applying these steps to our study, the following points outline how this Scoping review will be conducted:

1. Identify a Team
  a. The Team will consist of two medical students, both with an interest and prior knowledge in muscular-skeletal injuries in the sporting environment.
  b. The students will be guided and supervised by: Professor Lucy Hammond, a professor in Health Sciences Education; Assistant Professor Hollie White, an assistant-professor in health sciences.
2. Identify the Research Question
  a. There have been numerous studies that aim to direct the management of anterior process calcaneal fractures, but none found that are explicitly Scoping Reviews.
  b. The literature is limited with regards to this injury within the sporting environment. More guidance on this area would be useful to help manage patients that fall into the “athlete” population.
  c. Initial research into this field indicates that there is enough research to warrant a scoping review.
3. Identify Relevant Studies
  a. Similarly to a Systematic review, a search of resources such as PubMed can be used to help provide papers that outline how these fractures are best diagnosed and managed. Researchers will work together to draw together all potentially relevant papers.
  b. Additionally, there may also be value in including a wider base of literature in the review. If papers are limited on PubMed then a further search of Embase will be used.
  c. Keywords will likely include: anterior process; calcaneus; fractures.
  d. Papers will be found using the following search terms:
    i. Embase – exp calcaneus fracture/ or exp calcaneus/ or calcaneus.mp (and) anterior process (and) fracture.mp or fracture/
    ii. PubMed – calcaneus (and) anterior process (and) fracture
4. Select Studies to be included in the review
  a. The inclusion/exclusion criteria used for this study will be underpinned by the Population, Concept and Context (PCC) framework.
    i. Population: Adults between the ages of 18-65. This population is thought to be most representative of the athlete population. Papers that specifically include participants under 18 or over 65 will be excluded from this study due to the common physiological differences between these populations and the general adult population.
    ii. Concept: Management of anterior process of the calcaneus fractures.
    iii. Context: Adults with a diagnosis of an anterior process of the calcaneus fracture, looking to return to their baseline level of function, to represent a return to sport.
  b. Type of literature. Research papers will be primarily used. However, letters to editor and opinion pieces will also be selected if they meet the inclusion/exclusion criteria, if searches on PubMed/Embase are insufficient. It is felt that these often best represent the opinions of clinicians that are at the forefront of diagnosing and managing these injuries and so should be included in this study.
  c. Screening and data extraction software (e.g. Covidence) and reference management software (e.g. EndNote) will likely be required to help manage the papers that will be included.
  d. A minimum of two researchers will be used to decide whether a paper should be included in this study. Where there is unresolved disagreement, a third-party opinion will be sought from the project supervisor.
5. Chart the data
  a. Researchers will work together to design a charting form on Microsoft Excel. This form will set out what information it aims to draw from the selected studies. The form will contain headings such as: mechanism of injury; diagnostic tools;
  b. method of management; rehabilitation programmes; return to sport/function/outcome; positives of management option; negatives of management option; recovery time.
  c. Papers that do not provide the necessary information for every heading will not be excluded. As a scoping review, we want to draw as much information from as many valid and relevant studies as possible.
  d. Relevant extracts and data will be pulled from each of the papers that are selected for inclusion under the relevant heading, as listed above. Researchers will work independently during this stage, pulling extracts from the selected papers on to the charting form.
6. Collate, summarise and report the results.
  a. A narrative synthesis will be used to draw together the information from the charting form.
    i. It is expected that different papers will measure and assess competing management options differently.
    ii. As a scoping review, we will not attempt to quantify this data or formulate our own scoring system to rank the management options, as we believe this will be too subjective and could invalidate our findings.
    iii. However, we will analyse the breadth of available literature to help the reader understand the different management options available, collating and summarising the positives and negatives of each management option.
    iv. We will also look at how papers measure outcomes to be able to evaluate how outcomes could be assessed and measured for future studies, helping to guide future research.
7. Consult the stakeholders (optional)
  a. An orthopaedic consultant, who specialises in foot and ankle injuries, discussed this project with one of the researchers and highlighted the need for further and clearer guidance regarding this injury.
  b. It is the researchers aim to continue to liaise with the clinicians primarily responsible for diagnosing and managing these conditions, chiefly orthopaedic surgeons, throughout the process. This is to ensure that the aims of the study are met. Namely, to provide guidance to clinicians in managing these injuries.

### Data Use & Storage

This section is not entirely relevant to our proposed scoping review. We will be using public data taken straight from pre-existing journals. As the data is available in the public domain, we will not need to take any further measures to comply with the Data Protection Act and other regulations.

### Ethical Considerations

The benefit of this study is to highlight methods to provide more clarity and clinical guidance on how these specific fractures should be managed, particularly in athletes looking to return to sport. The hope is that these injuries, will be better managed with clearer guidance.

This Scoping Review will only be using data that is already publicly accessible. No personal data will be used and therefore no breaches in confidentiality are expected. This project will only be utilising data that is already available and is not collecting or generating further data from participants. As there are no participants involved in this study there is minimal physical risk, no restrictions of autonomy and no ethical permissions are being sought.

Neither the authors or Warwick University has received payments or services in the past 36 months from a third party that could be perceived to influence, or give the appearance of potentially influencing, the submitted work.

### Resources

End Note > Reference Management system

Covidence > Screening and Data extraction, for the management of records

Microsoft Excel & Word

PubMed

Embase

## Dissemination

The aim of the authors is to publish this study, making it publicly available. The study only draws upon information that is already publicly available and is not collecting or storing original data.

## Data Availability

All data produced in the present study are available upon reasonable request to the authors.

